# Polygenic risk score analysis suggests hypothyroidism as a risk factor for Alzheimer’s disease

**DOI:** 10.1101/2024.10.13.24315421

**Authors:** Ignazio S. Piras, Marcus A. Naymik, Janith Don, Nicholas J. Schork, Don Saner, Matthew J. Huentelman

## Abstract

Utilizing the All of Us cohort, this study explores the relationship between clinical conditions, and AD risk, shedding light on potential preventative measures. We selected a subset of non-AD affected individuals aged 60 and older (n = 72,044). Polygenic Risk Scores (PRS) were calculated to assess AD risk, which were then associated with medical conditions. A replication study was conducted in the UK Biobank to validate the findings. We found significant association between AD-PRS and nine conditions, with the most significant being hypothyroidism (adj-p < 1.3^-06^). The AD-PRS was significantly lower in participants with hypothyroidism, and was further confirmed in a replication study using the UK Biobank data (n=390,543; p= 1.3^-23^). Our results demonstrate that the absence of hypothyroidism might be a factor enhancing AD resilience since it can balance the genetic risk factors, as non-affected individuals exhibited a significantly higher AD-PRS than individuals with hypothyroidism.

## 1. Introduction

Alzheimer’s disease (AD) is a neurodegenerative disease characterized by a progressive decline in cognitive abilities and executive functions. This deterioration is associated with synaptic connection loss and neuronal death(Braak and Braak, 1991; Dubois et al., 2010). Two primary neuropathological markers of AD are the build-up of amyloid-β plaques (Aβ) and the presence of neurofibrillary tau tangles (NFTs)(Alzheimer’s Association, 2015). Among older adults, AD stands as the predominant cause of dementia. Current estimates suggest approximately 40 million individuals globally have AD, with this number expected to double every two decades, continuing at least up to 2050(Prince et al., 2015). Despite extensive research and clinical trials, only a few FDA-approved therapies exist for Alzheimer’s Disease (AD), offering minimal long-term benefits. Over the past decade, nearly 200 potential AD treatments have either failed to demonstrate clinical benefit or been discontinued(Yiannopoulou et al., 2019). Possible reasons for these clinical trial failures include late treatment initiation in AD progression, insufficient knowledge of AD’s complex pathophysiology, inadequate drug dosing, and incorrect or sub-optimal drug targets(Yiannopoulou et al., 2019).

Approximately, 70% of the risk for developing AD can be attributed to genetics(Ballard et al., 2011). However, modifiable risk factors significantly influence an individual’s risk profile as well. Presently, it is understood that addressing twelve known risk factors might prevent or delay up to 40% of dementia cases worldwide(Livingston et al., 2020). These modifiable risk factors include: low education, hypertension, hearing impairment, smoking, obesity, depression, physical inactivity, diabetes, low social contact, excessive alcohol consumption, traumatic brain injury, and air pollution. Other acquired factors include more generally cerebrovascular diseases(Mayeux and Stern, 2012), Type 2 Diabetes(Li et al., 2015), dyslypidemia(Popp et al., 2013), stress and inadequate sleep(Carroll et al., 2011; Proserpio et al., 2018). Conversely, resilience in AD (R-AD) is a phenomenon where individuals display the neuropathological, imaging, or fluid biomarker hallmarks of AD, but do not exhibit clinical signs of cognitive impairment(Seto et al., 2021). The identification and understanding of mechanisms underlying resilience represents an interesting therapeutic avenue, that could lead to the development of interventions enhancing or promoting resilience, thereby delaying or preventing the onset of AD(Neuner et al., 2022).

Giving the significance of risk and protective factors in the development of AD, we utilized the All of Us (AoU) cohort(Denny et al., 2019) to examine the relationship between clinical conditions and the risk of AD.

## 2. Materials and methods

### 2.1 “All of Us” project description

The All of Us (AoU) research program(Denny et al., 2019) aims to collect data from a diverse cohort of over a million participants from various regions of the U.S. The primary objective is to expedite biomedical research and enhance health outcomes, particularly for population groups historically underrepresented in biomedical research(Denny et al., 2019). Initiated in May 2018, the program currently enrolls participants 18 years of age or older from a network of more than 340 recruitment sites. Data collected includes: 1) health surveys (e.g., sociodemographic characteristics, overall health, lifestyle and medical history), 2) physical measurements (e.g., blood pressure, heart rate, weight, and height), 3) biospecimens (blood and urine samples), 4) electronic health records (e.g., medication history, laboratory results, and vital signs); 5) digital health information (e.g., Fitbit measurements), 6) bioassays (microarray and whole genome sequencing), 7) geospatial and environmental data. (e.g., weather, air quality, and pollutants). Further insights can be found in Denny et al.(Denny et al., 2019) and on the program portal (https://www.researchallofus.org/data-tools/workbench/).

### 2.2 Dataset selection

We selected participants from the AoU Controlled Tier Dataset v7 (CDRv7), which includes a total of 410,235 individuals, retrieving medical conditions (including medical examination). A subset of these samples is currently complemented by short-read Whole Genome Sequencing (srWGS) data (n = 240,319), that we used for the Polygenic Risk Score (PRS) computation. Specifically, we used the ACAF (allele count-allele frequency) which are stored as a PLINK format and were previously filtered using population-specific allele frequency (AF > 1%) or population-specific allele count (AC > 100) in any computed ancestry populations.

For the medical conditions analysis, assuming all participants were clinically assessed, we included individuals 60 years and older with associated srWGS data and from both sexes. The age was computed using the date of birth and the lower range of the CDR date (05/06/2018). Then, we selected control samples excluding individuals with at least one of the listed conditions or taking one of the medications associated with AD or dementia (Supplemental Table 1). Individuals with missing sex or date of birth were excluded from the study cohort. We defined this cohort as controls (CTL). After this first step, we obtained a dataset of 72,044 participants, including 21,938 unique medical conditions, distributed among 56,585 individuals. Average age of the participant was 69.0 ± 6.2, sex ratio F:M 1.27. The Alzheimer’s Disease (AD) sample was selected according to the ICD.9 code G30.9. Using the same demographic criteria as above, we were able to select a sample of 715 participants. Average age was 74.8 ± 7.51 years; sex ratio F:M: 1.17).

For the measurements, the number of participants varied depending on the specific variable, with a range from 2 to 78,265. The measurement maximum sample size did not align with the conditions because age estimation for those measurements was based on the date of testing, rather than on a fixed cutoff date. We report this information for each variable in the results tables. The Alzheimer’s Disease (AD) sample was selected according to the ICD.9 code G30.9.

### 2.3 AD Polygenic Risk Score calculation

AD Polygenic Risk Score (AD-PRS) was calculated as described by Marees et al.(Marees et al., 2018) using the SNPs from Lambert et al.(Lambert et al., 2013). We selected SNPs with False Discovery Rate (FDR) less than 0.05 (n = 3,799), and mapped to the Human Genome Reference Hg38. We were able to use 3,739 SNPs also included in AoU srWGS. The number of alleles present in each individual were counted and then multiplied by their corresponding GWAS β-value and finally summed across all SNPs for each individual’s PRS. Three groups were then selected by comparing the AoU cohort PRS to the TGEN GWAS case/control dataset, a collection of AD cases and controls there were both clinically characterized while living and subsequently neuropathologically assessed following death (Corneveaux et al., 2010): 1) AoU cohort with a PRS higher than the largest TGEN control (Higher Control); 2) AoU cohort with a PRS higher than the median of the TGEN cases (Above Median AD); 3) AoU cohort in the top PRS decile of scores (Top PRS).

### 2.4 Association of PRS with medical conditions

srWGS ACAF-filtered data for all the 72,044 controls and 715 AD patients were filtered for the 3,799 SNPs included in the AD-PRS using *PLINK v1.9* (Chang et al., 2015). After filtering, we retrieved 3,739 SNPs for the PRS computation (the 98.4% of the total). For each condition, we modeled a linear regression using the PRS as dependent variable, and the condition (presence/absence) as a predictor. Sex and age were included as covariates. The resulting model, implemented in R with the *lm* function was:

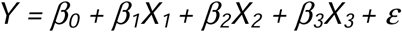

Where *Y* is the polygenic risk score, *X_1_* is the condition (yes = 1/ no = 0), *X_2_* sex, *X_3_* age, *β_0_* is the intercept of the model, *β_1_*, *β_2_*, and *β_3_* are the coefficients for the condition, sex and age, respectively. Finally, *ε* is the error term, accounting for the difference between the predicted and actual values of the outcome variable. Only conditions with a frequency of 1% or larger in our sample (n = 720) were used in the analysis. The results were adjusted for multiple testing using the Bonferroni correction, as implemented in the *multtest* R-package. The distribution of the top associated variables was further tested stratifying the controls samples according to the PRS percentile groups (Higher control, Above Median AD, and Top PRS; see “AD Polygenic Risk Score calculation” above for further details).

### 2.5 Association of PRS with measurements

srWGS ACAF-filtered data for all the 72,044 controls and 715 AD patients were filtered for the 3,799 SNPs included in the AD-PRS using *PLINK v1.9* (Chang et al., 2015). After filtering, we retrieved 3,739 SNPs for the PRS computation (98.4% of the total SNP panel that we targeted). Outliers were removed for each measurement using the interquartile range (IQR) method. Then, for each variable, we modeled a linear regression using the PRS as dependent variable, and the measurement as a continuous predictor. Sex and age were included as a covariate. The resulting model, implemented in R with the *lm* function was:

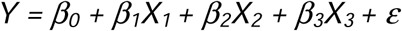

Where *Y* is the polygenic risk score, *X_1_* is the measurement (continuous variable), *X_2_* sex, *X_3_* age, *β_0_* is the intercept of the model, *β_1_*, *β_2_*, and *β_3_* are the coefficients for the measurement, sex and age, respectively. Finally, *ε* is the error term, accounting for the difference between the predicted and actual values of the outcome variable. Only measurements that comprise at least 1% of the sample (equivalent to 783 data points or more) were used in the analysis. The results were adjusted for multiple testing using the Bonferroni correction, as implemented in the *multtest* R-package. The distribution of the top associated variables was further tested stratifying the controls samples according to the PRS percentile groups (Higher control, Above Median AD, and Top PRS; see “AD Polygenic Risk Score calculation” above for further details).

### 2.6 Replication study in the UK Biobank

We replicated the results for the top associated variables in the UK Biobank dataset, including a total of 487,409 participants. After using the same filtering criteria as for the AoU sample (see above), we obtained a final UK Biobank cohort of 390,543 participants who underwent clinical assessment. Participants with hypothyroidism were defined according to the broad ICD10 codes categories “E02” and “E03”, and according to the broad ICD9 categories “243” and “244”. We used the UK Biobank imputed data version 3, genotyped with the array UK Biobank Axiom Array, and imputed using Haplotype Reference Consortium and UK10K + 1000 Genomes reference panels(Bycroft et al., 2017). After filtering the AD associated SNPs from Lambert et al.(Lambert et al., 2013) (n = 3,799), we obtained a total of 2,941 SNPs that we used to compute the AD-PRS as explained for the AoU cohort (see above). The association between AD-PRS and hypothyroidism prevalence was assessed using a liner model adjusted for age and sex.

## 3. Results

After including conditions with a frequency of 1% or larger in our sample (n ≥ 720), we were able to analyze 1,078 variables, finding significant results for nine conditions after Bonferroni adjustment (Supplemental Table 2, Figure 1). The top associated variables were hypothyroidism (β = −4.3; adj-p = 6.7^-11^) (Figure 2) and “acquired hypothyroidism” (β = −3.8; adj-p = 1.3^-06^) (Supplemental Fig S1). In both cases, the AD-PRS was significantly lower in participants diagnosed with the condition. We tested seven measurements variables associated with thyroid function, finding a significant association with autoimmune thyroiditis (β = −7.3; adj-p = 1.4^-04^) (Supplemental Table 3). Additionally, with the aim to investigate sex-specific differences, we tested the variables “hypothyroidism” and “acquired hypothyroidism” in both females and males, finding them in both cases as the top significant associated variables (Supplemental Table 4).

**Fig. 1.**
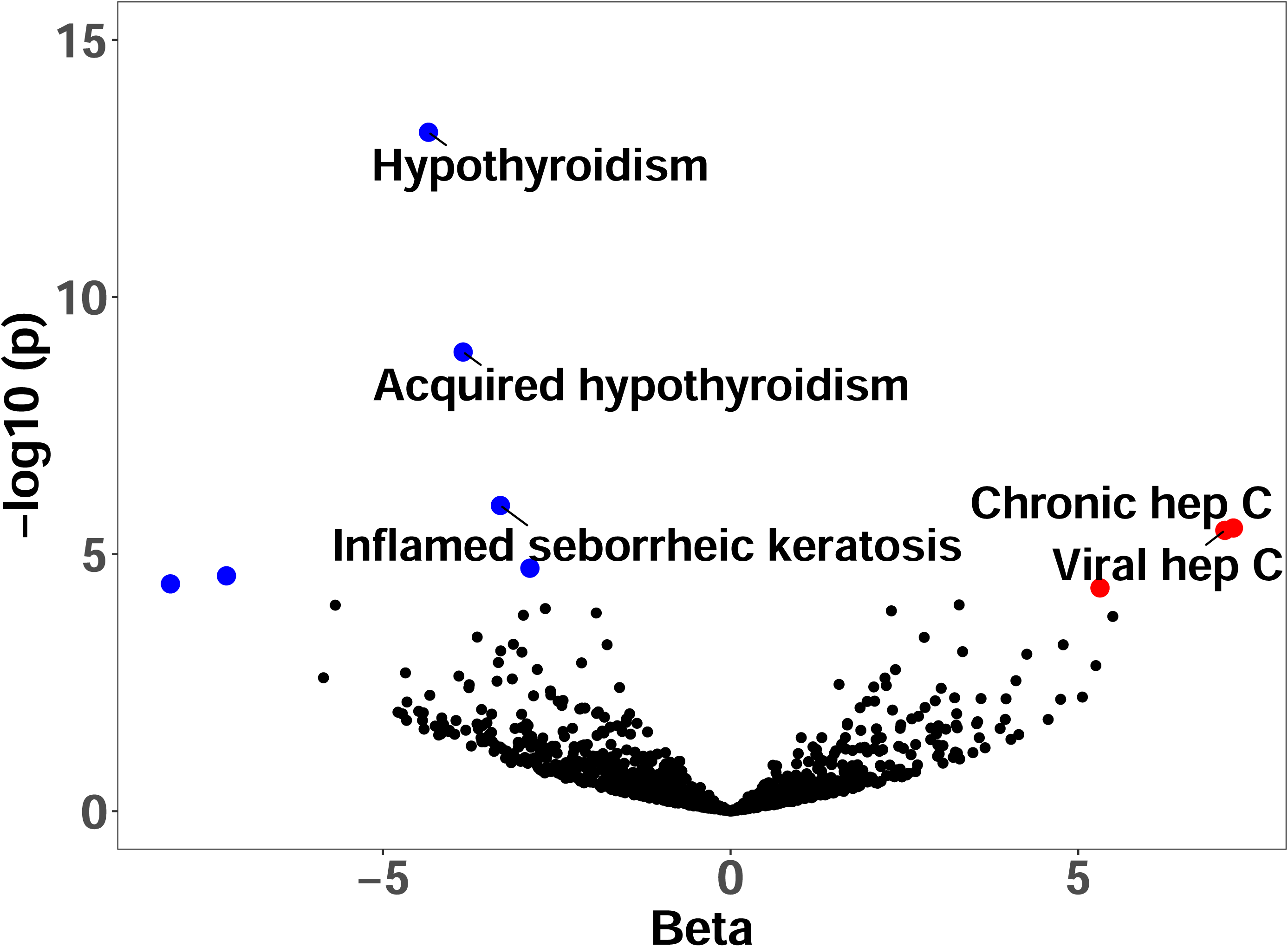
Volcano plot depicting the conditions associated with AD-PRS. The X-axis represents the regression coefficient, while the y-axis represents -log10 (adj-p)

**Fig. 2.**
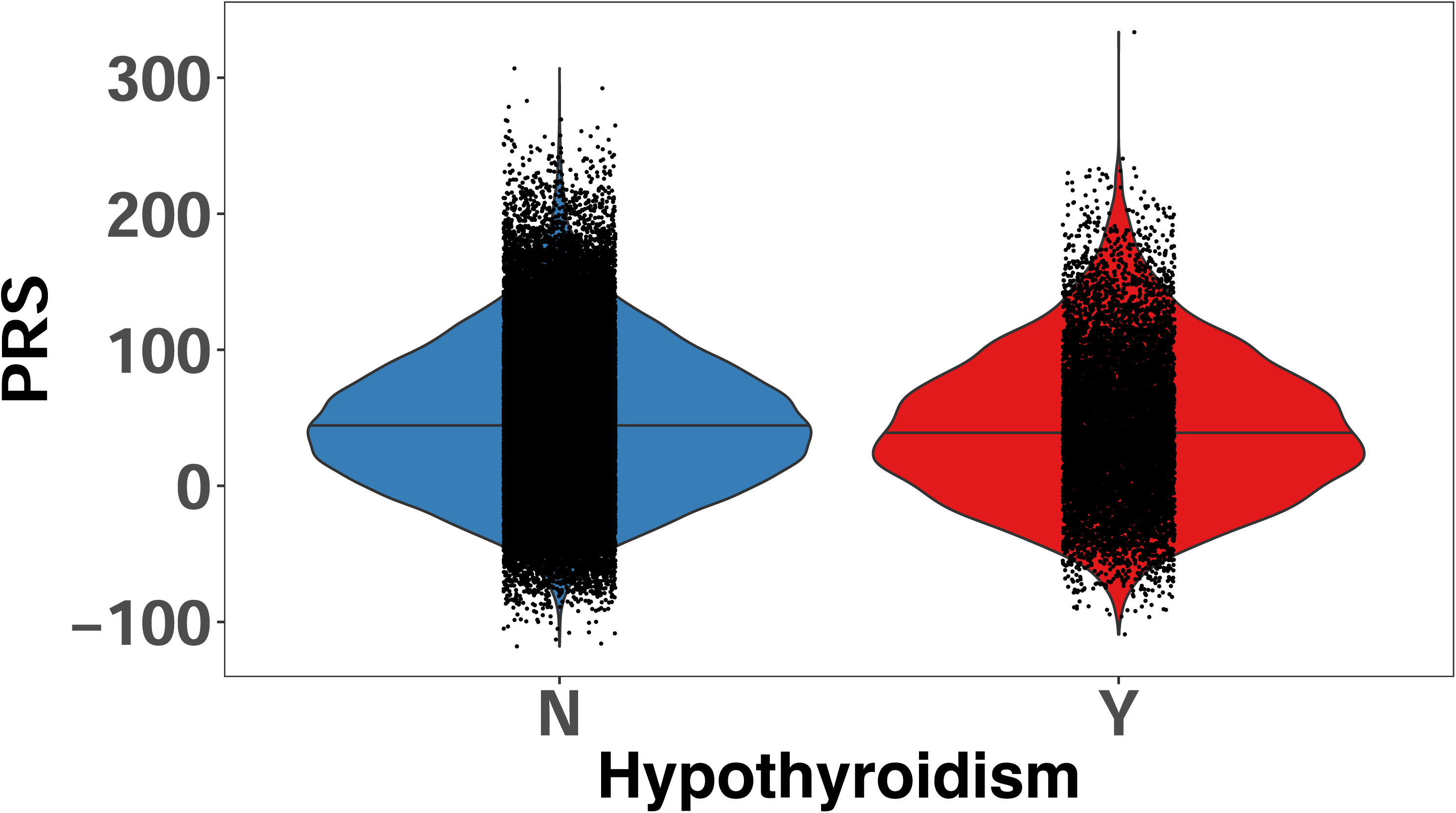
Violin plot illustrating the distribution of AD-PRS in participants diagnosed with hypothyroidism (Y) and non-affected individuals (N). The AD-PRS was significantly lower in affected individuals (β= −4.3; adj-p = 6.7^-11^).

Among other associated conditions, the association with inflamed seborrheic keratosis, melanocytic nevus, and splenomegaly followed the same trend, while the association with chronic hepatitis C, viral hepatitis C and talipes planus showed an increase of AD-PRS among participants with these conditions (Supplemental Table 2). We compared the proportion of hypothyroidism in AD (n = 715) versus the CTL, stratified by three different classes of AD-PRS (see methods for details). In all cases, the proportion of hypothyroidism was larger in AD than in all of the other three groups and this was statistically significant (Figure 3, Supplemental Table 5). We further investigated this association including the effect of hypothyroidism medications, taking into account the list of drugs in Table S6. After including the covariate “drug” in the linear model as a binary variable along with sex and age, the results remained significant for both hypothyroidism (β = −4.289; adj-p = 1.40^-13^) and acquired hypothyroidism (β = −3.377; p = 2.72^-09^). In both cases, the effect of the drug was not statistically significant (p > 0.081). Finally, we replicated these findings concerning the association between hypothyroidism and AD-PRS using the UK Biobank data (n=390,543), confirming the results after the adjustment for age and sex (β=-2.970; p=1.3^-23^) (Figure 4).

**Fig. 3.**
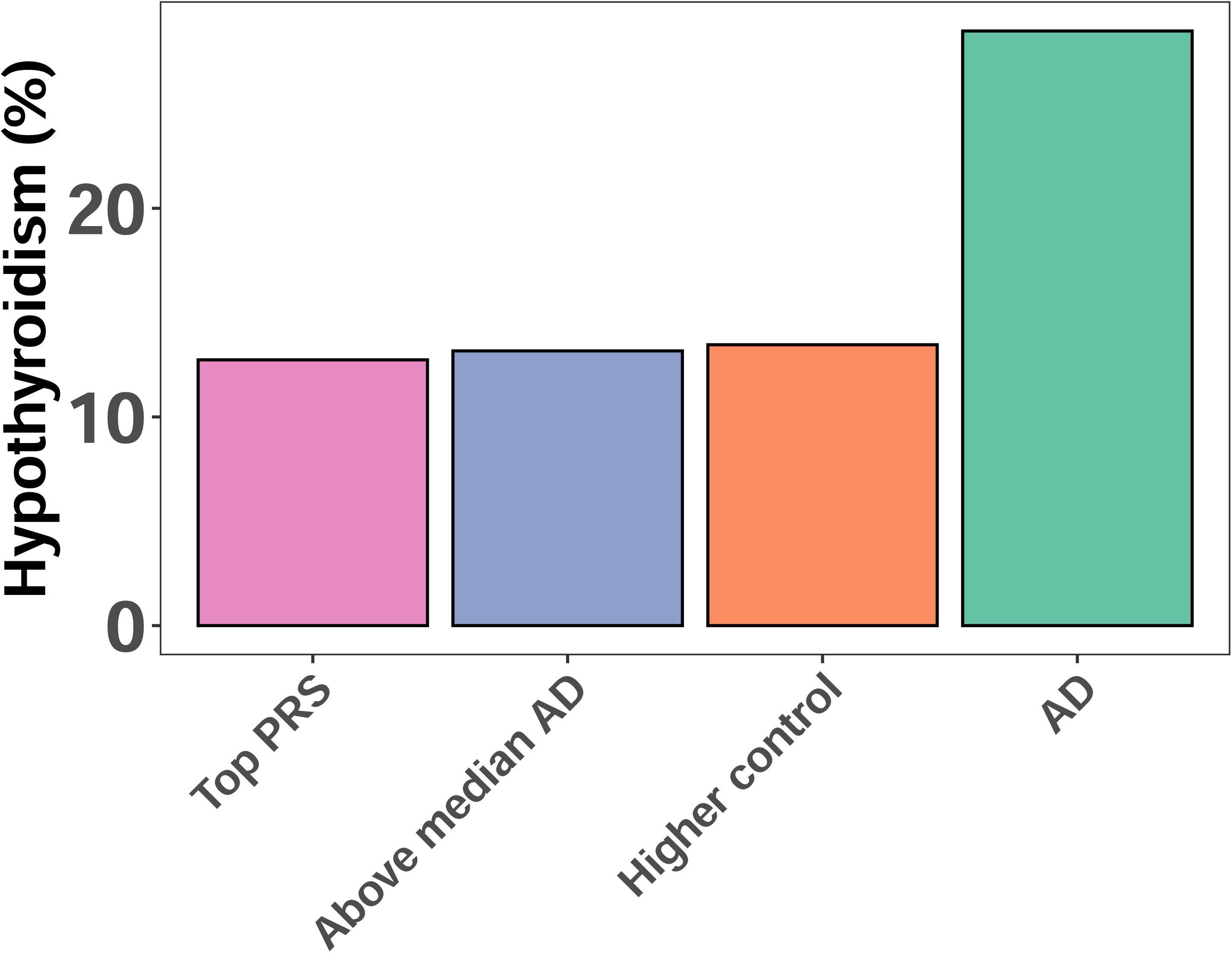
Comparison of the prevalence (%) of hypothyroidism in AoU cohort in AD and in the control group, stratified by AD-PRS score cutoffs. The prevalence was significantly higher in AD compared to all the three groups.

**Fig. 4.**
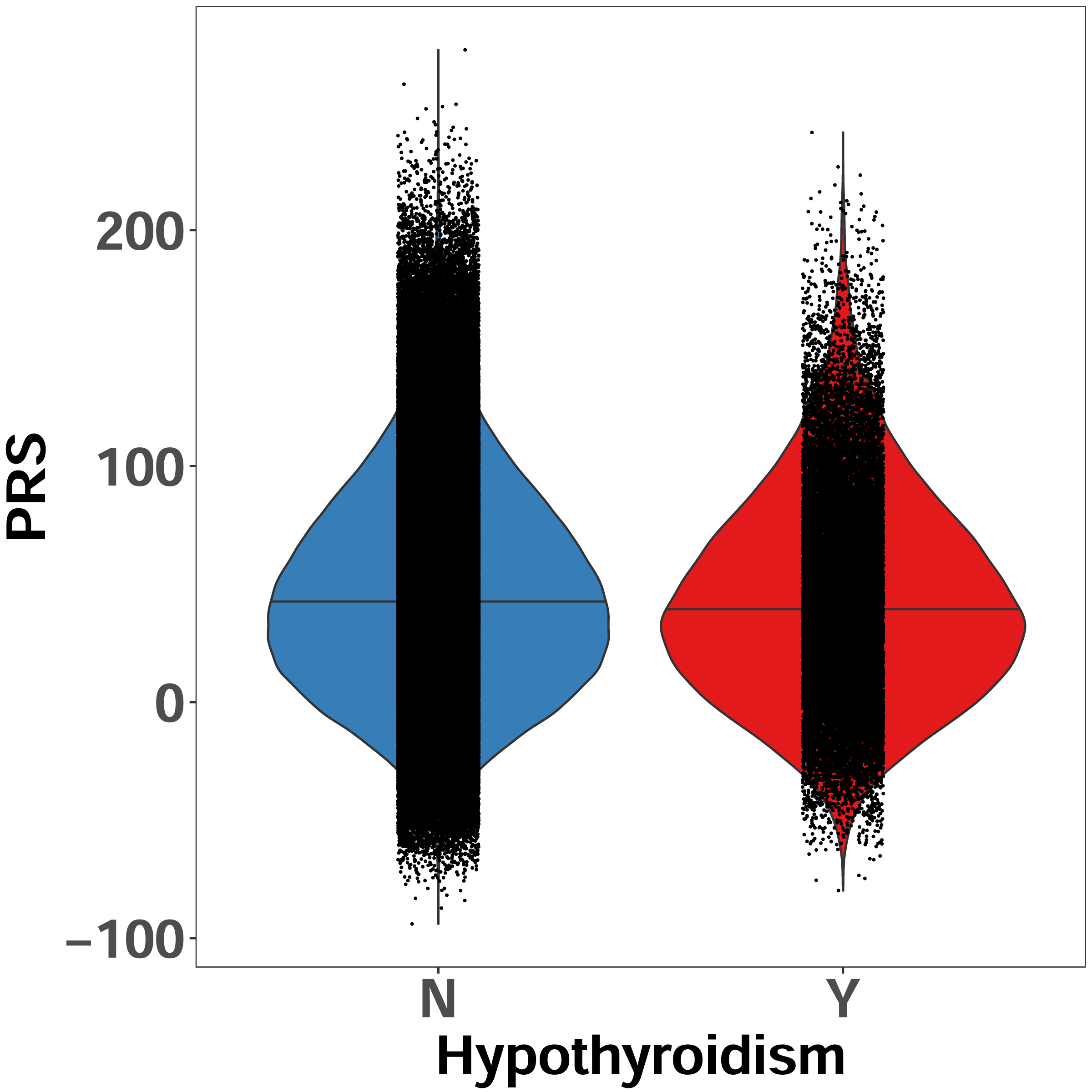
Replication of the hypothyroidism findings in the UK Biobank cohort. The AD-PRS was significantly lower in the participants diagnosed with hypothyroidism (β= −2.970; p=1.3^-23^).

## 4. Discussion

In this study, we investigated the association between various medical conditions and measurements with AD-PRS in a large cohort of older adult non-AD participants from the AoU cohort. Following strict quality control filtering, we analyzed the association of 1,078 conditions in more than 70,000 participants. Our goal was to identify variables associated with the risk of AD and/or resilience, that can be used for prevention strategies.

We identified significant associations between AD-PRS and several conditions after multiple testing correction. Notably, hypothyroidism, acquired hypothyroidism, and autoimmune thyroiditis emerged as top conditions with a statistically significant relationship with AD-PRS, without sex-specific effects. AD-PRS was significantly lower in individuals diagnosed with this condition. Additionally, autoimmune thyroiditis showed a significant association as well, with concordant direction (lower AD-PRS in affected). Interestingly, the thyroid-stimulating hormone level (TSH) exhibited a negative correlation with AD-PRS, reinforcing the results from the conditions analysis. TSH is produced by hypothalamus, and stimulates the thyroid to produce thyroxine and triiodothyronine. Levels of TSH higher than normal indicate hypothyroidism, since the thyroid gland is not producing enough hormones, and the hypothalamus increases the secretion of TSH to stimulate the thyroid. In our analysis, higher levels of TSH were associated with lower AD-PRS, which was associated with hypothyroidism in the conditions analysis. The association with hypothyroidism remained significant even when we included the hypothyroidism medications as a covariate. Additionally, the association between AD-PRS and hypothyroidism was successfully replicated in almost 400,000 samples using the genomics and clinical data from the UK Biobank. Finally, the prevalence of hypothyroidism in the AoU AD cohort was significantly higher than in the control sample. Our results demonstrate that the absence of hypothyroidism might be a factor enhancing resilience since it can balance the genetic risk factors, as non-affected individuals exhibited a significantly higher AD-PRS than hypothyroidism-affected individuals. Previous studies found an association between hypothyroidism, thyroid dysfunction, and AD risk (Salehipour et al., 2023; Wieland et al., 2022).

The relationship between AD and hypothyroidism seems to be bi-directional. One of the proposed mechanisms suggests that the neurodegeneration in AD leads to HPT, which enhances AD progression (AlAnazi et al., 2023). Several preclinical studies have demonstrated the effect of thyroid hormones on AD neuropathology (AlAnazi et al., 2023), such as the inhibition of *APP* gene expression, which leads to a reduction of amyloid plaque formation (Belakavadi et al., 2011), or HPT inducing the AB production in the hippocampus (Chaalal et al., 2014). Another link could involve the modulation of cerebral blood flow (Haji et al., 2015; Park et al., 2019). Reduction of T3 was associated with atrophy in brain cortex and hippocampus (Montero-Pedrazuela et al., 2006), whereas its administration (also T4) delays Ab pathology and neuroinflammation (Chaalal et al., 2019, 2014).

The most significant condition after hypothyroidism and acquired hypothyroidism was the inflamed seborrheic keratosis, with a lower AD-PRS in affected versus non-affected (β = −3.3; adj-p = 1.2E-03), similar to the observation in hypothyroidism. Inflamed seborrheic keratosis is a benign skin condition characterized by inflammation (Mansur and Yildiz, 2019; Roh et al., 2016). While there is no association between this condition and AD, it has been shown that APP and its downstream products were more highly expressed in the seborrheic keratosis tissue than in the adjacent normal skin tissue (Li et al., 2018). Additionally, it is known that skin physiology is altered in AD patients (Clos et al., 2012). We can speculate that lower AD-PRS in patients with seborrheic keratosis might increase resilience due to the chronic inflammation characteristic of the disease, associated with the increased risk of AD (Andronie-Cioara et al., 2023). Other significant top conditions were Chronic hepatitis C and Viral hepatitis C, with a opposite pattern compared to the hypothyroidism findings. Specifically, AD-PRS was higher in subjects with the condition. Hepatitis C is recognized as a risk factor for AD and dementia (Chiu et al., 2014). However, the causal relationship has not been assessed yet (Huang et al., 2022). The interpretation of our finding is not straightforward, but we can speculate about shared genetic risk factors between AD and hepatitis C, possibly related to the immune system function or inflammatory processes. To corroborate this hypothesis, a recent study found a shared genetic etiology between AD and circulating levels of 15 cytokines and growth factors(van der Linden et al., 2021). Our study includes participants with diverse ethnic background. This might limit the accuracy of PRS estimates, as the PRS was based using a GWAS with primarily European ancestry individuals.

In conclusion, our analysis revealed significant associations between AD-PRS and conditions like hypothyroidism, acquired hypothyroidism, and autoimmune thyroiditis. These conditions showed a lower AD-PRS, indicating potential resilience factors against AD. Thyroid-stimulating hormone levels also negatively correlated with AD-PRS, concordant with the other findings. Interestingly, inflamed seborrheic keratosis and hepatitis C showed opposite patterns, suggesting complex interactions between these conditions and AD risk, including, but not only, shared genetic etiology. These findings highlight potential options for AD prevention and underscore the need for further studies focused on the complex relationship between systemic diseases and neurodegeneration.

## CrediT taxonomy

**Ignazio S. Piras:** Data curation, Formal analysis, Methodology, Writing - original draft. **Marcus A Naymik:** Data curation, Formal analysis, Visualization. **Janith Don:** Validation, Visualization. **Nickolas J. Schork**: Methodology, Writing - review & editing. **Don Saner:** Methodology, Resources, Writing - review & editing. **Matthew J Huentelman**: Conceptualization, Investigation, Supervision, Writing - review & editing

## Consent statement

Informed consent was obtained from all the participants. All experimental protocols involving human participants were approved by Ethics committee/Institutional Review Board (IRB) of the All of Us Institutional Review Board

## Data Availability

All data analyzed herein were provided by All of Us research program and by UK Biobank. UK Biobank data were provided under project reference 43036.

https://www.researchallofus.org/data-tools/workbench/

https://www.ukbiobank.ac.uk/enable-your-research/about-our-data

