## Supplementary_figures for "Polygenic risk score analysis suggests hypothyroidism as a risk factor for Alzheimer’s disease"

**Supplemental Fig S1**. Violin plot showing the distribution of PRS across participants diagnosed with acquired hypothyroidism (Y) and non (N). AD-PRS was significantly lower in participants with acquired hypothyroidism (β = -3.8; adj-p = 1.3-06).

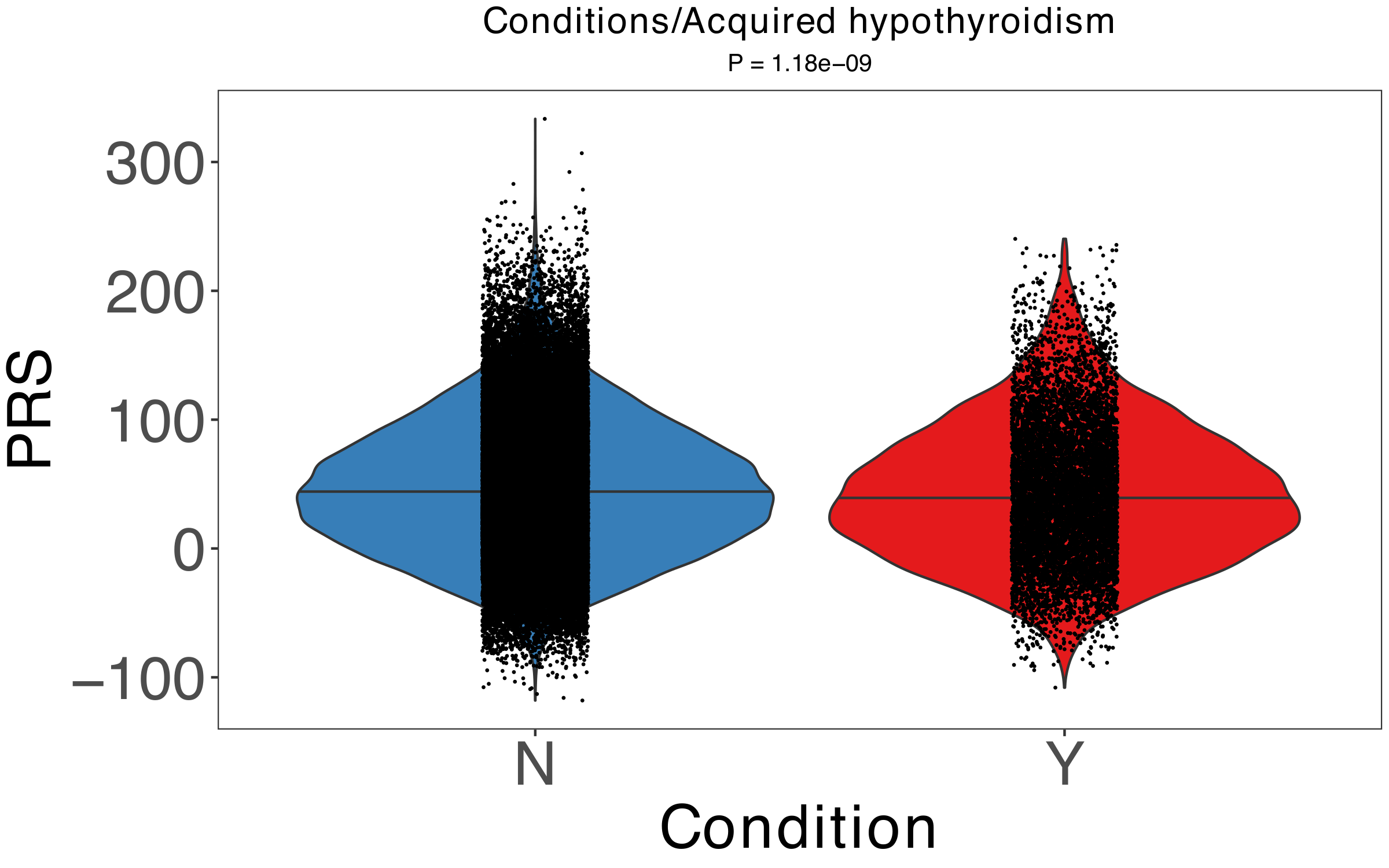
